# Feasibility and acceptability of collecting passive phone usage and sensor data via Apple SensorKit

**DOI:** 10.1101/2025.02.06.25321806

**Authors:** Courtney Funk, Zhuo Zhao, Adam G. Horwitz, Yu Fang, Karina Pereira-Lima, Vik Kheterpal, Srijan Sen, Elena Frank

**Affiliations:** Department of Psychiatry, University of Michigan Medical School, Ann Arbor, Michigan; Neuroscience Institute, University of Michigan, Ann Arbor, Michigan, United States of America; Department of Neurology, University of Michigan Medical School, Ann Arbor, Michigan, United States of America; CareEvolution, Ann Arbor, Michigan, United States of America

## Abstract

Privacy is a growing concern in mobile health research, particularly regarding passive data. Apple SensorKit provides a novel platform for collecting phone and wearable usage and sensor data, however the acceptability and feasibility of collecting these sensitive data to research subjects remains unknown. We piloted the SensorKit platform with a large sample of first-year U.S. medical residents as part of the longitudinal *Intern Health Study*. Findings demonstrate that successful enrollment and retention rates can be achieved in a longitudinal e-Cohort study that collects SensorKit data, however lower opt-in rates among racial minorities suggest the need for further evaluation of the equity implications around specific data types in mobile health research.

## Introduction

Digital phenotyping, a way to organize passively collected mobile technology data from participants in real time to better understand behavior, has the potential to utilize new technology and sensors to capture moment-to-moment behavior and better understand participant experiences [1,2]. Various studies have found that phone sensor data (e.g. GPS, accelerometer, light, or microphone) can capture circumscribed constructs of sleep, social context (e.g. relationships, social support), mood, and stress [3]. The availability of this data has enabled just-in-time adaptive interventions, which use conditions (e.g., location, physical activity, phone use) detected by passive sensors to deliver an intervention when it is expected to have the greatest impact [4,5].

In contrast to ecological momentary assessments, collection of this passive phone and sensor data places minimal burden on participants. However, studies have noted participant concerns related to privacy that may negatively impact participation [6,7]. Specifically, mHealth studies have found that a significant number of participants do not want to be monitored, tracked, or to provide private sensor-based data [8]. With the rapid expansion of mobile health technologies in recent years, digital health equity is also a growing concern [9]. However, most studies assessing the acceptability and privacy concerns have been small in scale, limiting sample generalizability and the assessment of specific subgroups including racial/ethnic minority groups [3,10,11].

Apple SensorKit provides a novel technological framework for health researchers to capture passive participant data in domains relevant to well-being. However, the acceptability of collecting this data among patients is not well-understood. To address this question, the Apple SensorKit platform was assessed with first-year medical residents in the United States via the MyDataHelps mobile application as part of the longitudinal *Intern Health Study* [12].

Incoming intern physicians for the 2023-2024 academic year were invited to enable the collection of Apple SensorKit data as an optional component during the process of initial enrollment and onboarding in the Intern Health Study, which took place on a rolling basis over a three month period in spring 2023. Five sensors were selected for inclusion based on potential utility for data analyses concerning the mental health and adaptive functioning in training physicians (Ambient Light, Keyboard Metrics, Message Usage, Phone Usage, and Frequently Visited Locations). SensorKit data collection was tracked as part of the study for the first two months of the intern year.

## Methods

### Ethics Statement

The study design was approved by the Institutional Review Board at the University of Michigan and the participating hospitals in the Intern Health Study (HUM00033029). All participants provided written informed consent via a secure online survey as approved by the University of Michigan IRB.

### Study Design

Between April 11, 2023 and June 30, 2023, incoming intern physicians for the 2023-2024 academic year were invited to opt into providing Apple SensorKit data. Five sensors were selected for inclusion based on potential utility for data analyses concerning the mental health and adaptive functioning in training physicians (Ambient Light, Keyboard Metrics, Message Usage, Phone Usage, and Frequently Visited Locations) (**Figure 1**).

**Figure 1.**
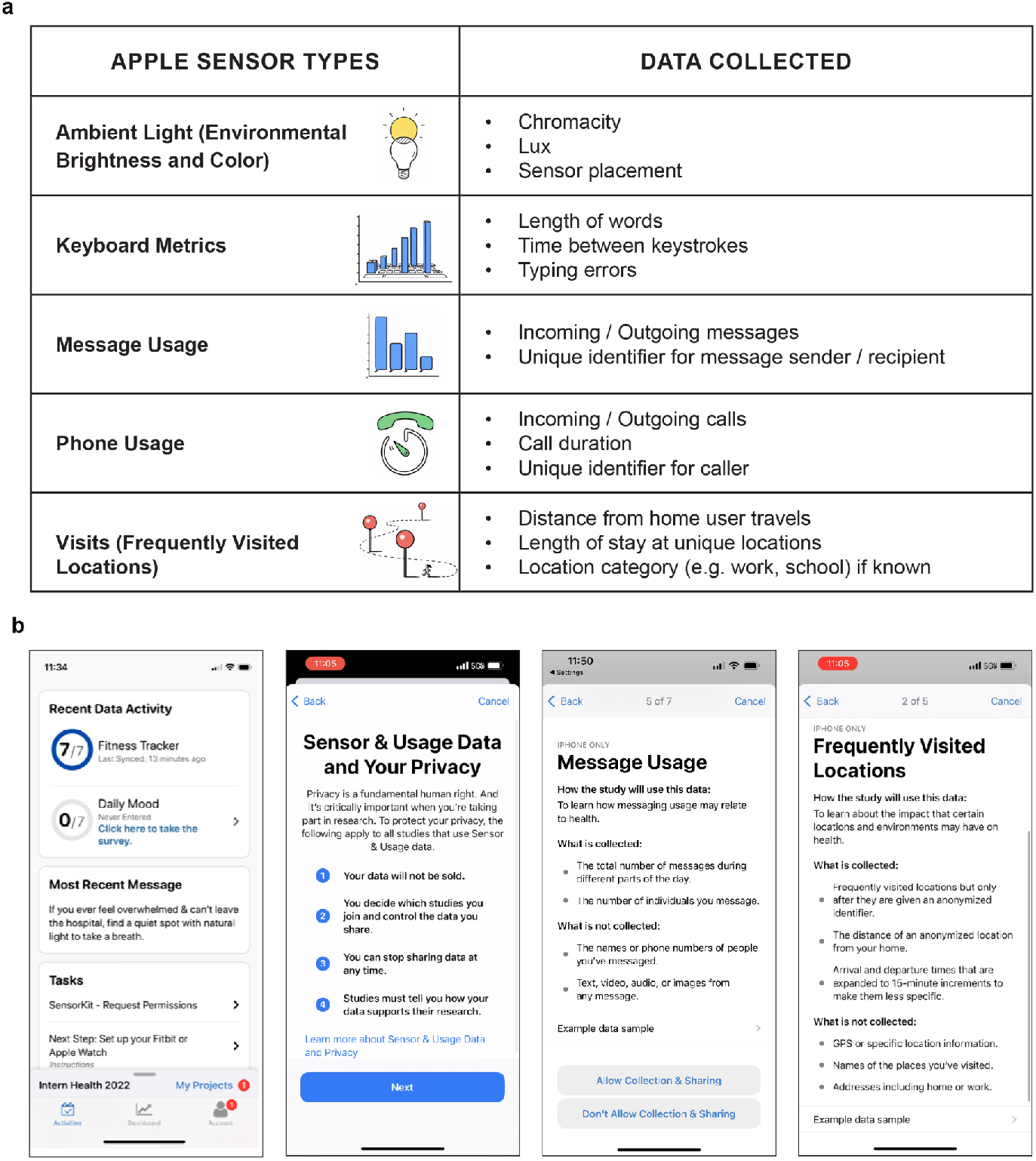
Apple SensorKit Types and Example Permission Screens. a Apple SensorKit data types selected for inclusion in the study. b Example screens of the participant opt-in process, including an overview of the requested data and two example data type description screens.

Participants completed consent and the initial survey of the parent study, and then logged into the MyDataHelps mobile app. The detailed Apple SensorKit Permissions screens appeared in the participant task list upon initial app login. Opening the permissions survey prompted participants with the standard Apple SensorKit introduction screen. Participants were then required to proceed through screens with detailed descriptions and examples for all sensors included in the present study (**Figure 1**). Following the information for each data type, participants were explicitly asked for permission to access that specific sensor: “Allow Collection & Sharing’’ or “Don’t Allow Collection & Sharing.” Participants could opt into data collection for all, some, or none of the sensor types presented. The data types were presented in alphabetical order as dictated by the Apple SensorKit platform. Participants were informed that enabling SensorKit was optional and could be disabled at any time. No additional incentives were provided for opting into the collection of SensorKit data. Device data were tracked as part of the study for the first two months of intern year.

### Data Analysis

We descriptively assessed the overall SensorKit opt-in and retention rates, and used chi-square analyses to assess differences in the initial opt-in rate and retention after 2 months between each of the 5 sensor options (Ambient Light, Keyboard Metrics, Messages Usage, Phone Usage, Frequently Visited Locations). We utilized chi-square analyses to evaluate differences in SensorKit enrollment based on the following demographic characteristics: age, gender, and race. For analysis purposes, racial groups were combined into the following categories: White, Asian, and underrepresented in medicine (URiM) (**Table 1**). Residents were coded as URiM according to the American Association of Medical Colleges definition as “racial and ethnic populations that are underrepresented in the medical profession relative to their numbers in the general population”24. In this study, this group included interns self-identifying as African American, Arab or Middle Eastern, Latino, Native American, Pacific Islander, other, or multi-racial. We then conducted chi-square analyses to compare national intern race data provided by the American Association of Medical Colleges (AAMC)12 with enrollment in the parent study in order to assess the overall racial representativeness of the sample. Analyses were conducted using SAS version 9.4 (SAS Institute). Statistical tests were 2-sided and used a significance threshold of P<.05.

**Table 1.** Demographic Characteristics (n=695)

| Gender | n (%) |
| --- | --- |
| Women | 396 (57.0) |
| Men | 298 (42.9) |
| Missing | 1 (0.1) |
| Mean age (IQR), y | 27.7 (26-29) |
| Ethnicity |  |
| White | 399 (57.4) |
| Asian | 136 (19.6) |
| Underrepresented in medicine<sup>a</sup> | 159 (22.9) |
| Missing | 1 (0.1) |
| Specialty<sup>b</sup> |  |
| Surgical | 133 (19.1) |
| Non-surgical | 562 (80.9) |

<sup>a</sup>Residents were coded as underrepresented in medicine according to the American Association of Medical Colleges definition as "racial and ethnic populations that are underrepresented in the medical profession relative to their numbers in the general population." In this study, this group included interns self-identifying as African American, Arab or Middle Eastern, Latino, Native American, Pacific Islander, other, or multi-racial.
<sup>b</sup>Surgical specialties were assigned based on the American College of Surgeons classification. Specifically, for this study, physicians in the following specialties were classified as "surgical": General Surgery, Gynecology and Obstetrics, Neurological Surgery, Orthopaedic Surgery, Otolaryngology, Plastic Surgery, Urology, and Other surgical. Physicians from the following specialties were classified as "non-surgical": Internal Medicine, Pediatrics, Psychiatry, Neurology, Emergency Medicine, Internal Medicine-Pediatrics, Family Medicine, Family Practice, Anesthesiology, Dermatology, Medical Genetics, Nuclear Medicine, Pathology, Physical Medicine & Rehabilitation, Preventative Medicine, Radiation Oncology, Radiology-Diagnostic, Sleep Medicine, and Other Non-surgical.

## Results

Among the 1437 incoming intern physicians who enrolled in the parent study, 1164 were iPhone users and invited to participate in the SensorKit arm of the study. Of those, 695 (59.7%) enabled at least one SensorKit data type during initial study onboarding. A significant difference in opt-in rates was observed between data types at enrollment (*P<*.*001, ES=0*.*08)* and at 2 months (*P<*.*001, ES=0*.*08)*, with Ambient Light as the highest enrolled and Frequently Visited Locations as the lowest enrolled at both time points **(Figure 2)**. At 2 months, 94% (653/695) of participants were still providing SensorKit data. **Figure 2. Apple SensorKit Enrollment and Retention Rates at Month 2 by Data Type**

**Figure 2.**
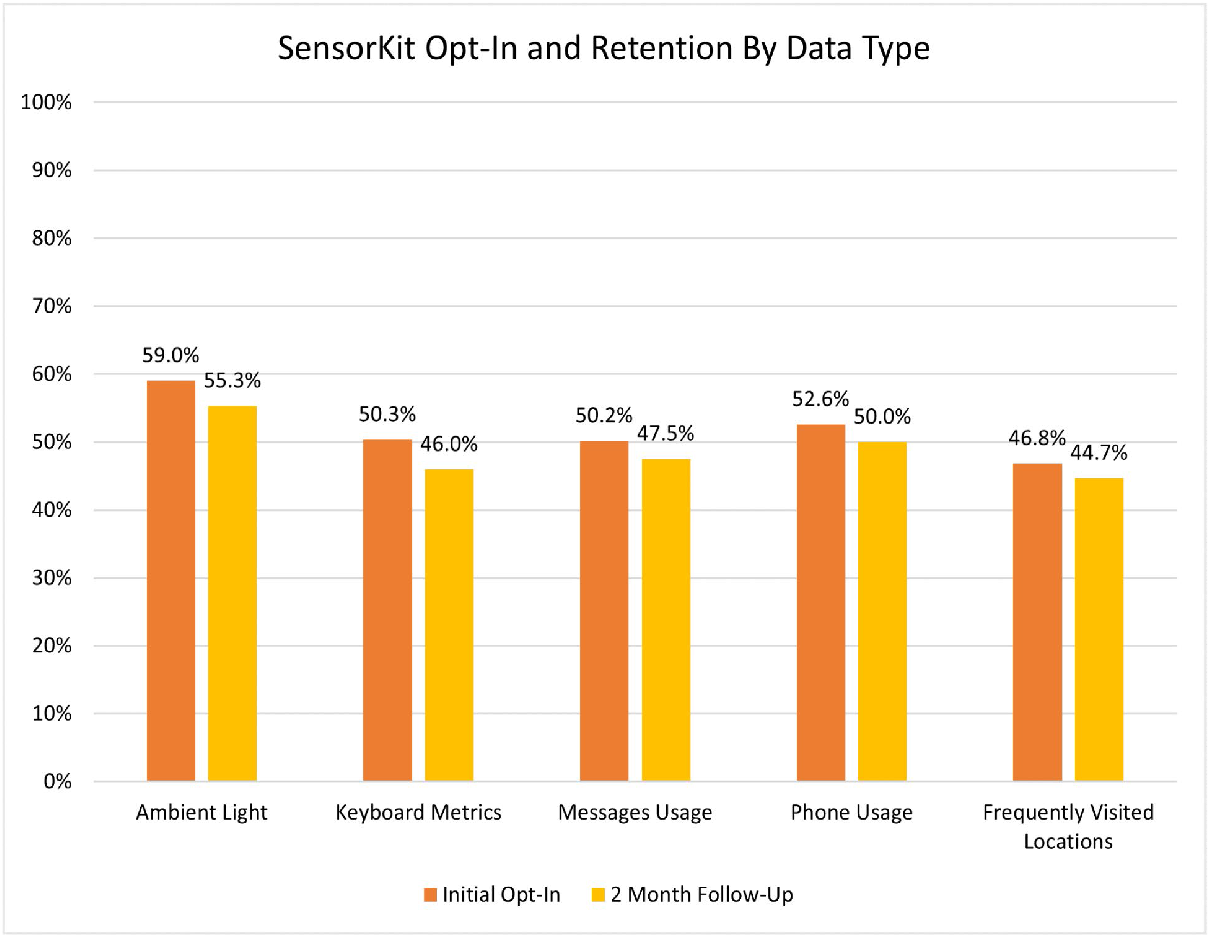
Apple SensorKit Enrollment and Retention Rates at Month 2 by Data Type

There were no significant differences in opt-in rates based on age or gender. However, interns who identified as White (55.1% [399/724]) were significantly more likely to opt into providing SensorKit data than those who identified as Asian (42.9% [136/317]) or as races underrepresented in medicine (43.6% [159/365] (*P<*.*001, ES=0*.*12*)). We utilized national intern data provided by the American Association of Medical Colleges (AAMC) [13] to determine if the parent study enrollment rate also differed by race and found no significant difference; the racial difference only emerged at the SensorKit opt-in level.

## Discussion

Data privacy and security are a growing concern in mobile health research [14,15].Yet, little is known about the feasibility of collecting this data on a large scale, or its acceptability by research participants [16]. In this study, we establish that successful participation rates can be achieved in a carefully designed longitudinal e-Cohort study that includes the collection of Apple SensorKit data. However, this may be contingent on SensorKit opt-in remaining optional. Future research should investigate how requiring participants to enable particular SensorKit data types affects enrollment rates.

Further, we found that the opt-in rate for Ambient Light was significantly greater than the other data types, with the lowest opt-in rate for Frequently Visited Locations (geographical location). Even though it is clearly stated in the SensorKit permissions screens that GPS data and specific addresses are not collected, it is possible that it was not clear to participants how this differed from other applications that do collect and exploit this kind of personal information. It is also unclear whether this difference was due to variation in privacy concerns with each data type, or because the data types were presented in alphabetical order as required by Apple, with the highest opt-in rate for the sensor presented first (“Ambient Light”) and the lowest rate for the sensor presented last(“Visits”). Further investigation of the role of sensor type order on opt-in rates is needed.

Additionally, once SensorKit is enabled, we find that the overwhelming majority of participants (93.8%) continue to provide SensorKit data even after 2 months. This stands in contrast to adherence to daily mood reports in the Intern Health Study, which often declines over the course of the year, and highlights the advantage of passive sensing for longer-term monitoring and detection [17]. Furthermore, while most recent studies on sensor technologies do not focus on user engagement and adherence, a few mHealth studies have found poor study adherence, reporting close to fifty percent of participants uninstalling their app or stopping engagement in the app within weeks of enrollment [8,18]. In the future, SensorKit data could be analyzed with corresponding daily-level mood data to identify predictive features– allowing for mood inferences to be made strictly by ongoing SensorKit data long after participants stop completing daily self-report mood ratings.

While the proportion of each racial group enrolled in the overall Intern Health Study closely reflected national US intern data, we found that interns identifying as Asian or as races/ethnicities underrepresented in medicine (e.g., African American, Latino) were significantly less likely to opt into providing SensorKit data than their peers identifying as White. Most research on the “digital divide” has focused primarily on age and socioeconomic disparities [19, 20]. However, this work indicates that there may be an important racial difference in acceptability of digital phenotyping, consistent with racial differences in other classes of medical research due to historical discrimination [20,22]. This aligns with qualitative work that suggests that experiences of discrimination and surveillance from employers and government institutions may lead to increased awareness of privacy risks among marginalized groups, and greater likelihood of taking steps to protect online privacy [23]. Similarly, a recent study of social media users found higher reports of privacy concerns among Asian and Latinx users compared to White users [24].

Much of the research in this area suggests that the primary driver of a potential “online privacy divide” is lower socioeconomic status and fewer resources (e.g. access to technology skills training) [25]. Yet, this study shows that even among a population of young people with iPhones, and the same educational and occupational status, racial identity still affects willingness to opt in to potentially more sensitive sensor and phone usage data. These findings suggest that motivation for privacy management in this population may be based more on identity and associated experiences (e.g. discrimination) rather than inequalities in resources, which may lead to lack of trust.

Beyond privacy concerns, other recent work suggests the potential for racial bias in mobile health research, based on potential inaccuracies in wearable device data for people with darker skin tones [26]. Prior concerns about representation in research including smartphone sensor data have also been raised due to variability in smartphone penetration across different global regions, low participation rates in mHealth studies, and possible differences between smartphone owners and non-owners [27]. Future studies should more closely investigate drivers of these disparities and consider implications for mobile health equity more broadly.

## Limitations

To our knowledge, this study is the first to evaluate SensorKit enrollment and retention rates in a national e-Cohort Study. Our large sample size allowed us to assess differences in the acceptability of various data types, and identify the potential for racial bias in the collection of sensor and phone usage data more generally. One limitation of the current study is the sample homogeneity in terms of age and education level, limiting the generalizability of the results. Participants in their late twenties with a medical degree may be more comfortable with technology or have fewer privacy concerns compared with older populations or individuals with other educational backgrounds. Another limitation was our inability to customize, and ideally randomize, the sensor type order during the opt-in process due to the technical constraints of the Apple SensorKit platform. This may have affected overall willingness to participate (for example, if a more “sensitive” sensor type was presented early on), or resulted in fewer participants enabling data types presented later. As the SensorKit data collection platform is only available for Apple devices, we were unable to include participants without iPhones in this analysis.

Future research should assess enrollment and retention rates for the collection of comparable sensor data in other smartphone types, as well as assess the accuracy and reliability of sensor data collected using various platforms. Using qualitative methods to further investigate potential barriers to participation, particularly as relates to such issues as privacy and security, could also be useful.

## Conclusion

Taken together, passive sensing methods such as those in SensorKit provide an opportunity to detect and intervene with individuals struggling with mental or physical health problems, particularly those that are chronic and have extended periods of potential risk. While there is significant potential for these methods, combined with computational advancements, to improve health outcomes, these opportunities must be balanced against privacy concerns and individuals’ willingness to participate in these types of programs. Significant health disparities already exist for minoritized populations and mHealth approaches must be cautious not to extend this gap.

## Data Availability

All data produced in the present study are available upon reasonable request to the authors

## Acknowledgements

We thank John Brussolo and Siqing Hu from the University of Michigan HITS database team for their support of this project.

